# Pregnancy and risk of COVID-19

**DOI:** 10.1101/2021.03.22.21254090

**Authors:** Maria C. Magnus, Laura Oakley, Håkon K. Gjessing, Olof Stephansson, Hilde M. Engjom, Ferenc Macsali, Petur B. Juliusson, Anne-Marie Nybo Andersen, Siri E. Håberg

## Abstract

**Background:** Studies report that pregnant women with coronavirus disease 2019 (COVID-19) are at increased risk of severe disease, intensive-care and death. Whether pregnant women in general are more susceptible of contracting severe acute respiratory syndrome coronavirus 2 (SARS-CoV-2) is unclear.

**Methods:** Linked registry-data on all women ages 15 to 45 living in Norway on March 1^st^, 2020 (N=1,033,699) were used in Cox regression models to estimate hazard ratios (HR) comparing pregnant to non-pregnant women, of having a positive test for SARS-CoV-2, a diagnosis of COVID-19 in specialist healthcare, or hospitalization with COVID-19, adjusting for age, marital status, education, income, country of birth and underlying medical conditions.

**Results:** Compared to non-pregnant women, pregnant women had a similar risk of a positive SARS-CoV-2 test (adjusted HR, 0.99; 95% confidence interval [CI], 0.92 to 1.07), a higher risk of a COVID-19 diagnosis in specialist care (HR, 3.46; 95% CI, 2.89 to 4.14), and to be hospitalized (HR, 4.70; 95% CI, 3.51 to 6.30). Pregnant women were in general not more likely to be tested for SARS-CoV-2. Pregnant women born outside Scandinavia were less likely to be tested, but at higher risk of a positive test (HR, 2.37; 95% CI, 2.51 to 8.87) and of hospitalization with COVID-19 (HR, 4.72; 95% CI, 2.51 to 8.87) than pregnant Scandinavian born women.

**Conclusion:** Pregnant women were not more likely to be infected with SARS-CoV-2. However, pregnant women with COVID-19, especially those born outside of Scandinavia, were more likely to receive specialist care and to be hospitalized.

---

It is unclear if pregnant women have an increased risk of severe acute respiratory syndrome coronavirus 2 (SARS-CoV-2) infection, but emerging evidence suggest that pregnant women are at higher risk of severe coronavirus disease 2019 (COVID-19) if infected.^1-4^ Most existing studies were from single centers or on hospitalized women with COVID-19, and investigated whether pregnancy increased the risk of severe disease, admission to intensive-care units, mechanical ventilation, and death.^5,6^ Population-based estimates comparing pregnant women compared to non-pregnant women are lacking.

The aim of this study was to examine whether pregnant women in general were at a higher risk of SARS-CoV-2 infection, or to be in contact with specialist healthcare services for COVID-19, compared to non-pregnant women, using data from national health-registries on all women in Norway between 15 and 45 years of age.

## Methods

### Study population and data sources

We followed all women between 15 and 45 years of age registered in the Norwegian National Population Registry on March 1^st^, 2020 (n= 1,033,699), until February 28^th^, 2021. Information on pregnancies and antenatal care visits was obtained from the birth registry, the patient registry (covering specialist/secondary healthcare services), and the general practitioner database (covering general practitioners/primary healthcare services).^7^ Information on SARS-CoV-2 tests was provided from the Norwegian Surveillance System for Communicable Diseases, while contacts with specialist healthcare services for suspected and confirmed COVID-19 were obtained from the patient registry. Information on education (highest level attained as of 2019) and household income (in 2018) was from Statistics

Norway. Data was linked by using unique personal identification numbers. Ethical approval was obtained from the Regional Committee for Medical and Health Research Ethics of South/East Norway (# 141135). Data from all registries was provided by the Emergency Preparedness Register for COVID-19 at the Norwegian Institute of Public Health.^8^ More information on data sources is in the Supplementary Appendix.

### Definition of completed pregnancies

The birth registry provided data on live births, stillbirths, fetal losses and induced abortions after 12 gestational weeks. Registrations of miscarriages and induced abortions occurring before 12 gestational weeks were obtained from the patient registry and the general practitioner database, as previously described.^9^ The diagnostic codes used to define miscarriage and induced abortion are in Table S1 in the Supplementary Appendix. These early miscarriages and induced abortions do not have registrations on gestational length of the pregnancy. Based on the mean gestational length for all induced abortions in Norway in the anonymous abortion registry, and the gestational age distribution of miscarriages from the literature,^10^ we assigned these pregnancies a gestational duration of 8 weeks, and in sensitivity analyses a gestational duration of 6 weeks or 10 weeks.

### Definition of ongoing pregnancies

We identified ongoing pregnancies using codes for antenatal care visits in the general practitioner database and the patient registry (see Table S2 in the Supplementary Appendix). These antenatal codes capture virtually all pregnancies that eventually will be recorded in the birth registry, as 99.5% of pregnancies in the birth registry had at least one registration of these codes during pregnancy. For a pregnancy to be defined as “ongoing” at the end of the study period, we excluded registrations occurring within the duration of a completed pregnancy. Second, we required that registrations of the antenatal codes were at least 90 days after a completed pregnancy to be counted as a new/currently ongoing pregnancy. Antenatal codes are not registered with a gestational length. Based on the distribution of the first registration of an antenatal code for the already completed pregnancies in the birth registry (Figure S1 in the Supplementary Appendix), we defined the start date of ongoing pregnancies to be 5 weeks (35 days) before the first antenatal consultation, assuming that very few women have an antenatal visit before 5 weeks of pregnancy. In additional analyses we assigned these pregnancies to start 10 weeks before the first visit.

### COVID-19

We defined COVID-19 in three ways: 1) a positive test for SARS-CoV-2, 2) any diagnosis of COVID-19 in specialist healthcare, and 3) hospitalization with confirmed COVID-19. Two new ICD-10 codes were implemented at the start of the pandemic: U07.1 “COVID-19 with confirmed virus”; and U07.2 “COVID-19 without confirmed virus”. These codes were used to define specialist diagnosed COVID-19. We further analyzed hospitalization for confirmed COVID-19 (U07.1) separately.

### Pre-existing chronic conditions

We obtained information on a wide range of pre-existing chronic condition defined as risk factors for severe COVID-19.^11^ The diagnostic codes we used to define these conditions are shown in Table S3 in the Supplementary Appendix. We required at least two registrations from January 2017 until end of follow-up to qualify as an existing underlying condition.

### Statistical analysis

We used Cox proportional hazards models on calendar time to examine separately whether pregnant women had an increased risk of 1) a positive test; 2) a specialist care diagnosis of COVID-19; and 3) hospitalization with confirmed COVID-19. Women were followed from March 1^st^, 2020, until the event of interest; emigration, death, or reaching February 28^th^, 2021 without an event was treated as censoring. Pregnancy status was a time-varying exposure, allowing women to contribute both pregnant and non-pregnant follow-up time. We used robust cluster variance estimation with the woman‘s identification number as the cluster variable. We estimated unadjusted associations, and associations with adjustment for marital status (single, married/cohabitating, or other), educational level (elementary school, high-school, vocational, up to 4 years of higher education, and more than 4 years of higher education), household income (categorized into tertiles), country of birth (Scandinavian countries (Norway, Sweden and Denmark) or non-Scandinavian countries), and chronic conditions. We first analyzed the entire follow-up period, and subsequently analyzed the two main waves of the pandemic in Norway separately ^12^ (March 1^st^ to June 30^th^, 2020, and July 1^st^ 2020 to February 28^th^ 2021). We also evaluated if associations differed with pregnancy trimester (1^st^ trimester: ≤83 days; 2^nd^ trimester: 84-195; and 3^rd^trimester: ≥196 days). As a higher risk of COVID-19 has been reported among non-Scandinavian ethnic groups in Norway,^13^ we also examined the risk of COVID-19 separately for Scandinavian and non-Scandinavian born women.

It could be that pregnant women were tested more often, and that milder COVID-19 therefore was detected more often among pregnant women resulting in higher estimates of COVID-19 among pregnant women. We examined whether pregnant women were tested more often than non-pregnant women. Women could have multiple tests during follow-up. We used the Andersen and Gill recurrent events Cox model,^14^ where women continued to be a part of the risk set until emigration, death or end of follow-up. To evaluate whether testing in relation to admission to hospital for delivery or miscarriage/abortion was driving the associations, we performed sub-analyses where we excluded tests conducted within three days before or after a pregnancy ended, and in addition hospitalizations where the end of pregnancy happened within a hospital stay for COVID-19. All analyses were conducted in Stata version 16 (Statacorp, Texas).

## Results

Of the 1,033,699 women included in the study, 101,820 (10%) had been pregnant during the follow-up time. There were 35,915 (4%) who were still pregnant at the end of follow-up (ongoing pregnancies). There was a slightly higher proportion of women born outside of Scandinavia among the pregnant women than among non-pregnant women (Table 1). Fewer pregnant women had chronic underlying risk conditions (Table 1).

**Table 1.** Distribution of characteristics among 1,033,699 ages 15 to 45 in Norway who were pregnant between March 1^st^, 2020 and February 28^th^, 2021.

| **Characteristics** | **Women who were pregnant**<br>**(n=102,820)** | **Women who were not pregnant**<br>**(n=930,879)** |
| --- | --- | --- |
| **Age at start of follow-up, mean (SD)** | 30.8 (5.1) | 30.2 (8.8) |
| **Country of birth, no. (%)** |  |  |
| Norway | 73,936 (71.9) | 705,553 (75.8) |
| Another Scandinavian country | 2,026 (2.0) | 14,186 (1.5) |
| Outside of Scandinavia | 26,528 (25.8) | 208,193 (22.4) |
| Unknown | 330 (0.3) | 2,947 (0.3) |
| **Marital status, no. (%)** |  |  |
| Single | 59,163 (57.5) | 636,473 (68.4) |
| Married/registered partner | 39,520 (38.4) | 241,090 (25.9) |
| Other | 4,137 (4.0) | 53,316 (5.7) |
| **Educational level, no. (%)** |  |  |
| Elementary school | 16,243 (15.8) | 221,684 (23.8) |
| Highschool | 19,416 (18.9) | 230,053 (24.7) |
| Vocational | 1,566 (1.5) | 13,955 (1.5) |
| Up to 4 years of university | 37,289 (36.3) | 272,101 (29.2) |
| More than 4 years of university | 20,049 (19.5) | 102,543 (11.0) |
| Unknown | 8,257 (8.0) | 90,543 (9.7) |
| **Household income, no. (%)** |  |  |
| 1<sup>st</sup> tertile (≤ 500,730 NOK) | 30,241 (29.4) | 304,914 (32.8) |
| 2<sup>nd</sup> tertile (500,731 to 846,668 NOK) | 41,219 (40.1) | 293,937 (31.6) |
| 3<sup>rd</sup> tertile ($>$ 846,668 NOK) | 28,081 (27.3) | 307,073 (33.0) |
| **Unknown** | **3,279 (3.2)** | **24,955 (2.7)** |
| **Chronic conditions, no. (%)** |  |  |
| **Diabetes** | **1,203 (1.2)** | **10,365 (1.1)** |
| **Cerebrovascular disease** | **104 (0.1)** | **1,339 (0.1)** |
| **Other chronic cardiovascular disorders** | **823 (0.8)** | **6,783 (0.7)** |
| **Immune deficiency** | **37 (0.04)** | **453 (0.05)** |
| **Reduced immune function due to medications** | **1,566 (1.5)** | **14,713 (1.6)** |
| **Chronic lung disease** | **3,505 (3.4)** | **36,953 (4.0)** |
| **Neurological disorders** | **93 (0.1)** | **2,263 (0.2)** |
| **Kidney failure** | **27 (0.03)** | **507 (0.05)** |
| **Organ transplant** | **21 (0.02)** | **628 (0.07)** |
| **Hematological cancer** | **95 (0.1)** | **1,036 (0.1)** |
| **Other types of cancer** | **94 (0.1)** | **2,405 (0.3)** |

### Risk of a positive SARS-CoV-2 test

The overall rate of a positive SARS-CoV-2 test among women aged 15-45 years was 5 per 100,000 person-days. The risk of a positive test was similar for pregnant women and non-pregnant women; adjusted HR, 0.99; 95% CI, 0.92 to 1.07, with similar HRs across all trimesters (Table 2). The estimate was similar for the two waves of the pandemic (first wave, adjusted HR, 0.94; 95% CI, 0.76 to 1.17, and second wave, adjusted HR, 1.00; 95% CI, 0.92 to 1.08; Table 2). Results were also similar after excluding women with positive tests within three days around the end of pregnancy (Table S4 in the Supplementary Appendix). Women born outside of Scandinavia had an increased risk of a positive test compared to Scandinavian women in general, and even higher risk when pregnant; adjusted HR, 2.37; 95% CI, 1.98 to 2.84, when compared to Scandinavian pregnant women (Table S5 in the Supplementary Appendix).

**Table 2.** Hazard ratio of a positive SARS-CoV-2 test during pregnancy among 1,033,698^*^ women in Norway between 15 and 45 years of age.

| Follow-up period | Pregnancy status | Follow-up time in days | No. of positive tests | Hazard Ratio (95% CI) |  |
| --- | --- | --- | --- | --- | --- |
|  |  |  |  | Without Adjustment | With Adjustment<sup>†</sup> |
| **Complete follow-up<sup>‡</sup>** | Non-pregnant | 356,383,248 | 16,364 | 1.00 | 1.00 |
|  | Pregnant | 15,481,516 | 708 | 0.98 (0.91-1.05) | 0.99 (0.92-1.07) |
|  | 1st trimester | 5,454,096 | 256 | 0.97 (0.86-1.10) | 0.98 (0.87-1.11) |
|  | 2nd trimester | 5,787,833 | 271 | 0.99 (0.88-1.12) | 1.01 (0.90-1.14) |
|  | 3rd trimester | 4,239,587 | 181 | 0.96 (0.83-1.11) | 0.97 (0.84-1.13) |
| **Wave 1<sup>§</sup>** | Non-pregnant | 119,435,417 | 1977 | 1.00 | 1.00 |
|  | Pregnant | 5,198,569 | 87 | 1.01 (0.82-1.26) | 0.94 (0.76-1.17) |
|  | 1st trimester | 1,746,753 | 24 | 0.87 (0.58-1.30) | 0.81 (0.54-1.21) |
|  | 2nd trimester | 1,941,362 | 35 | 1.05 (0.75-1.46) | 0.97 (0.69-1.36) |
|  | 3rd trimester | 1,510,454 | 28 | 1.13 (0.78-1.65) | 1.05 (0.72-1.53) |
| **Wave 2** | Non-pregnant | 236,947,831 | 14,387 | 1.00 | 1.00 |
|  | Pregnant | 10,282,947 | 621 | 0.97 (0.90-1.05) | 1.00 (0.92-1.08) |
|  | 1st trimester | 3,707,343 | 232 | 0.98 (0.86-1.12) | 1.01 (0.88-1.15) |
|  | 2nd trimester | 3,846,471 | 236 | 0.99 (0.87-1.12) | 1.02 (0.90-1.16) |
|  | 3rd trimester | 2,729,133 | 153 | 0.93 (0.79-1.09) | 0.96 (0.82-1.12) |

\*Excluded one person that tested positive before March 1<sup>st</sup>, 2020.
†Adjusted for age as a linear and squared term, country of birth, marital status, education, household income, diabetes, cerebrovascular disease, other cardiovascular disorders, immune-deficiency, chronic lung disease, reduced immune function, neurological disorders, kidney failure, organ transplant, hematological cancer, and other types of cancer.
‡March 1<sup>st</sup>, 2020 to February 28<sup>th</sup>, 2021
§March 1<sup>st</sup>, 2020 to June 30<sup>th</sup>, 2020
||July 1<sup>st</sup>, 2020 to February 28<sup>th</sup>, 2021

### Risk of specialist-care diagnosis and hospitalization

The overall rate of a specialist healthcare diagnosis of COVID-19 was 0.3 per 100,000 person days, while the rate of being hospitalized with confirmed COVID-19 was 0.1 per 100,000 person days. Pregnant women had an increased risk of a specialist care diagnosis of COVID-19 (adjusted HR, 3.46; 95% CI, 2.89 to 4.14), which was similar in both waves of the pandemic (Table 3). The risk appeared to be highest in the third trimester but was attenuated when we excluded pregnancies ending within the same hospital stay as for COVID-19 (Table 3). The increased risk of contact with specialist healthcare services for COVID-19 while pregnant were higher in non-Scandinavian pregnant women (adjusted HR, 7.50; 95% CI, 5.76 to 9.77), and Scandinavian pregnant women (adjusted HR, 2.66; 95% CI, 2.09 to 3.39), when compared to Scandinavian women who were not pregnant (Table S6 in the Supplementary Appendix).

**Table 3.**
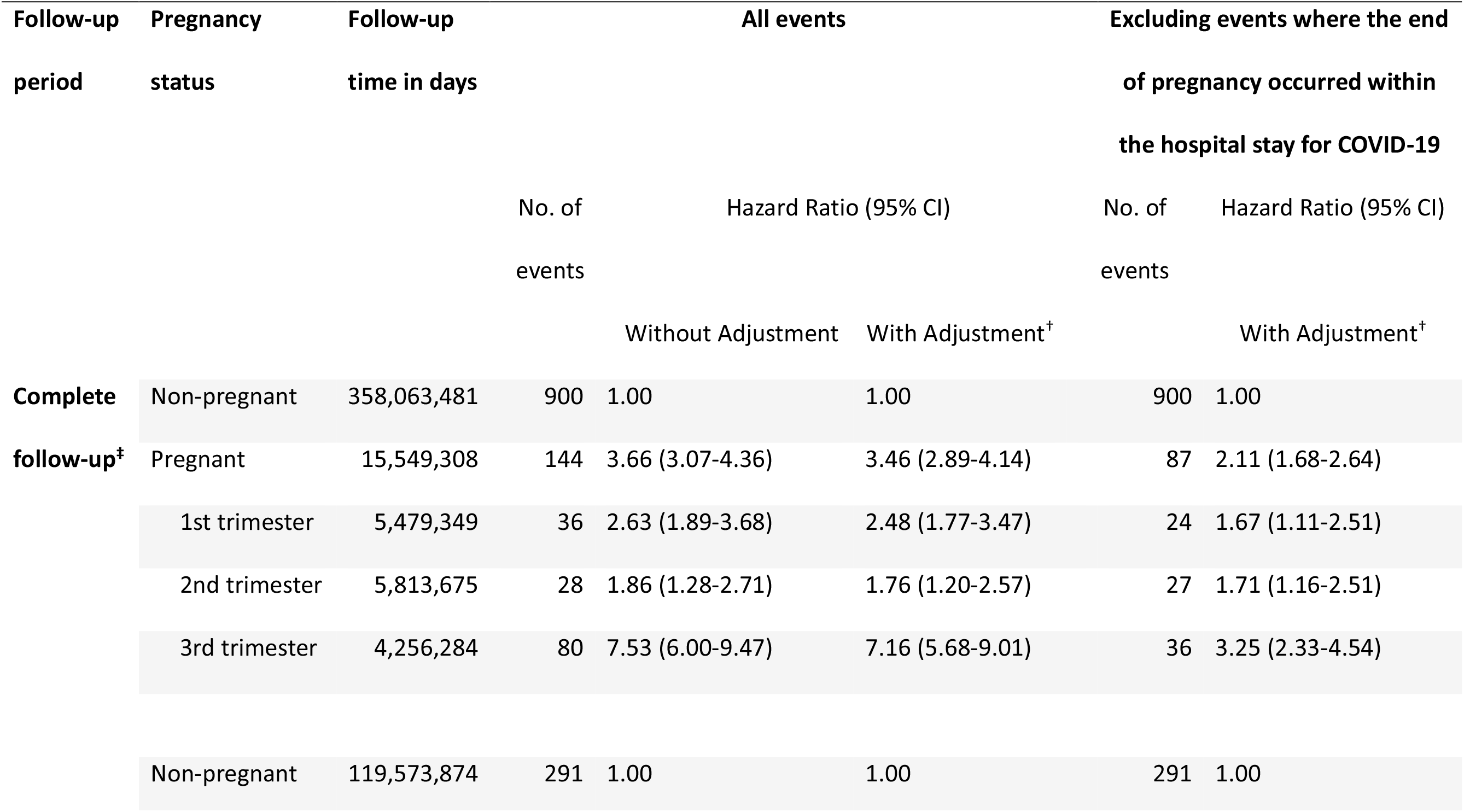
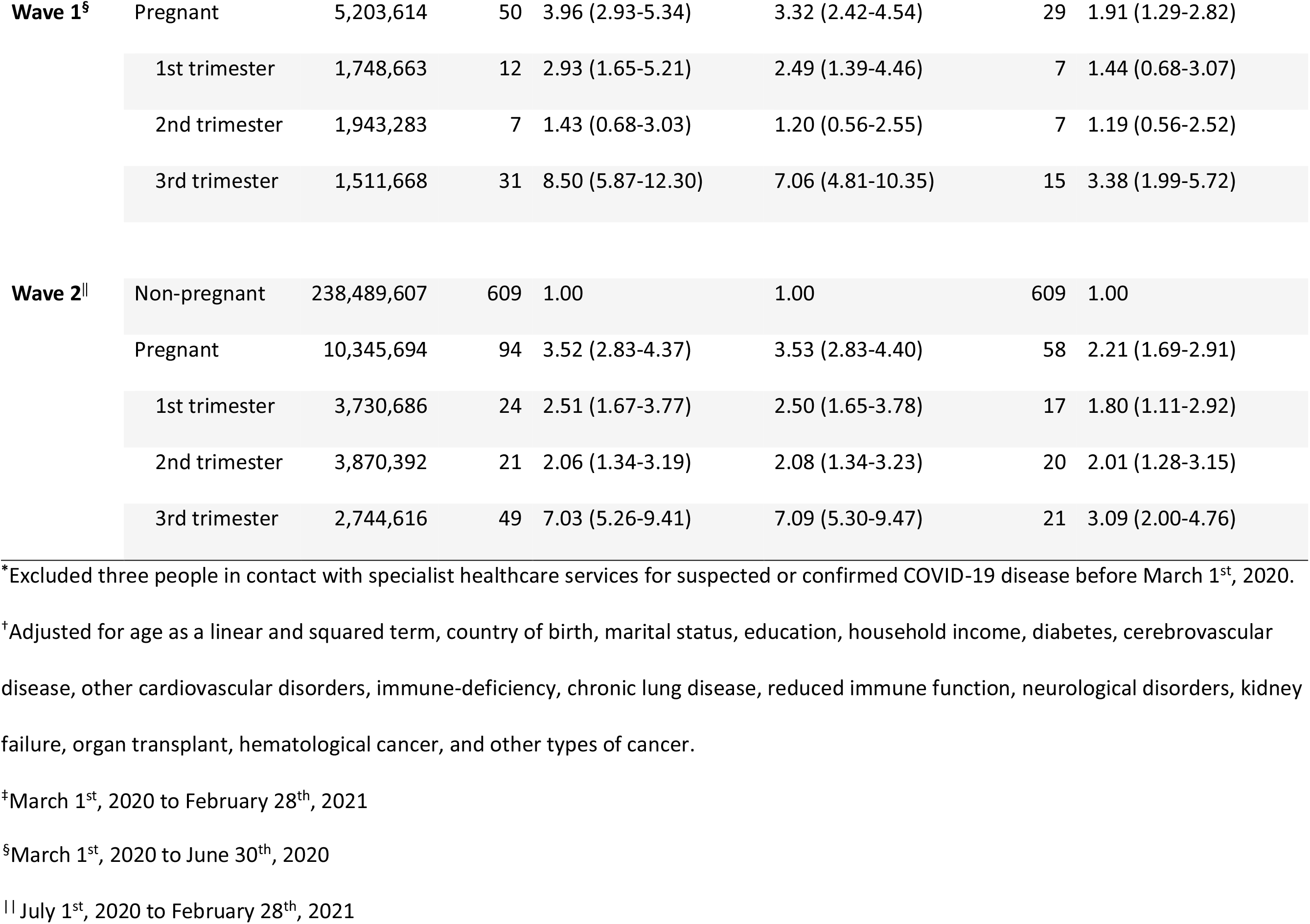
Hazard ratio of a COVID-19 diagnosis in specialist healthcare services for pregnant women among 1,033,696^*^ women between 15 and 45 years of age in Norway.

Pregnant women had a substantially higher risk of being hospitalized for confirmed COVID-19, adjusted HR, 4.70; 95% CI, 3.51 to 6.30, in both waves of the pandemic (Table 4). The greatest risk was seen in the third trimester, though the trimester-specific differences were attenuated when we excluded pregnancies ending within the same hospital stay where COVID-19 was diagnosed. Among COVID-19 hospitalized women, the proportion who also had diagnoses of lower respiratory illness (ICD-10 codes J12-J22, J80, J96) was 32% in pregnant and 49% in non-pregnant women. The median number of days in hospital was 2 for pregnant (mean 3.3 days) and 2 for non-pregnant women (mean 3.7 days).

**Table 4.** Hazard Ratio of Hospitalization (Event) with Confirmed COVID-19 for Pregnant Women among 1,033,699 Women Between 15 and 45 Years of Age.

| Follow-up period | Pregnancy status | Follow-up time in days | All Events |  |  | Excluding events where the end of pregnancy occurred within the hospital stay for COVID-19 |  |
| --- | --- | --- | --- | --- | --- | --- | --- |
|  |  |  | No. of events | Hazard Ratio (95% CI) |  | No. of events | Hazard Ratio (95% CI) |
|  |  |  |  | Without Adjustment | With Adjustment* |  | With Adjustment* |
| Complete follow-up<sup>†</sup> | Non-pregnant | 358,173,181 | 289 | 1.00 | 1.00 | 289 | 1.00 |
|  | Pregnant | 15,559,886 | 53 | 4.19 (3.12-5.61) | 4.70 (3.51-6.30) | 24 | 2.21 (1.45-3.37) |
|  | 1st trimester | 5,482,901 | 8 | 1.81 (0.89-3.66) | 2.00 (0.99-4.06) | 6 | 1.55 (0.69-3.49) |
|  | 2nd trimester | 5,817,698 | 11 | 2.27 (1.25-4.15) | 2.58 (1.41-4.72) | 10 | 2.44 (1.30-4.59) |
|  | 3rd trimester | 4,259,287 | 34 | 10.01 (7.01-14.27) | 11.37 (7.97-16.21) | 8 | 2.78 (1.37-5.65) |
| Wave 1<sup>‡</sup> | Non-pregnant | 119,591,018 | 88 | 1.00 | 1.00 | 88 | 1.00 |
|  | Pregnant | 5,205,118 | 15 | 3.93 (2.27-6.80) | 4.17 (2.37-7.31) | 6 | 1.70 (0.73-3.97) |
| **Wave 2<sup>§</sup>** |  |  |  |  |  |  |  |
|  | Non-pregnant | 238,582,163 | 201 | 1.00 | 1.00 | 201 | 1.00 |
|  | Pregnant | 10,354,768 | 38 | 4.30 (3.04-6.08) | 4.96 (3.52-6.98) | 18 | 2.45 (1.51-3.98) |

\*Adjusted for age as a linear and squared term, country of birth, marital status, education, household income, diabetes, cerebrovascular disease, other cardiovascular disorders, immune-deficiency, chronic lung disease, reduced immune function, neurological disorders, kidney failure, organ transplant, hematological cancer, and other types of cancer.
<sup>†</sup>March 1<sup>st</sup>, 2020 to February 28<sup>th</sup>, 2021
<sup>‡</sup>March 1<sup>st</sup>, 2020 to June 30<sup>th</sup>, 2020
<sup>§</sup>July 1<sup>st</sup>, 2020 to February 28<sup>th</sup>, 2021

Both being pregnant and being non-Scandinavian increased the risk of hospitalization with confirmed COVID-19, and pregnant non-Scandinavian women were at highest risk of hospitalization with COVID-19 (Table S7 in the Supplementary Appendix).

### Likelihood of being tested for SARS-CoV-2

The SARS-CoV-2 testing rate was 310 tests per 100,000 person days. Overall, pregnant women were slightly less likely to be tested for SARS-CoV-2, adjusted HR, 0.90;95% CI, 0.88 to 0.91 (Table S8 in the Supplementary appendix). The rate of testing in pregnant compared to non-pregnant women has been similar or lower after the initial pandemic months (Figure S2 in the Supplementary appendix). Lowest test rates among pregnant women were seen during third trimester (Table S8 in the Supplementary appendix). Non-Scandinavian women had lower probability of testing, especially when pregnant, adjusted HR, 0.72; 95% CI, 0.70 to 0.74, compared to non-pregnant Scandinavian women (Table S9 in the Supplementary appendix).

In additional analyses we reassigned the gestational duration of pregnancies ending in miscarriages and induced abortions to be 6 and 10 weeks, and ongoing pregnancies to start 10 weeks prior to first antenatal visit instead of 5 weeks; the results were very similar to the main analyses.

## Discussion

We found no overall increased risk of a positive SARS-CoV-2 test among pregnant women compared to non-pregnant women. However, pregnant women were at a substantially increased risk of requiring specialist healthcare and hospitalization.

Women born outside of Scandinavia were less likely to be tested, and at a particularly higher risk of being hospitalized for COVID-19 when pregnant compared to Scandinavian born women. An increased risk of COVID-19 among ethnic minorities has been reported in several countries,^15,16^ including Norway.^12^ This has been attributed to crowded households and more service related professions with personal contact. We observed a less testing among both pregnant and non-pregnant women born outside of Scandinavia. A higher threshold for testing may have resulted in more severe illness before seeking healthcare, which is supported by our findings of increased risk of specialist care and hospitalizations than Scandinavian born women. Routine testing of minority women in connection with antenatal care could reduce these differences.

This study is unique in its size as it included all women of reproductive age in Norway, with the ability to compare the pregnant with the non-pregnant population of similar age. We were also able to examine whether differences in testing behavior were likely to influence results, which was not found to be the case.

A limitation of registry studies is that health definitions rely on registrations from contact with healthcare. Test capacity for SARS-CoV-2 and healthcare availability for those with milder COVID-19 symptoms have varied through the pandemic. In the initial phase, testing was limited, and healthcare was restricted to those with more severe symptoms or underlying risk conditions. We found that pregnant women were slightly more likely to be tested in the initial phase than non-pregnant women, but after the initial months when testing capacity increased, pregnant women were slightly less likely to be tested.

Asymptomatic women and women with milder symptoms were not captured and classified as infected in our data. However, results for the two waves of the pandemic in Norway yielded similar estimates, supporting that test-or healthcare availability was unlikely to explain our findings.

Identifying ongoing pregnancies and early terminations through healthcare contacts is also prone to misclassification. Towards the end of the follow up period we were less likely to capture ongoing pregnancies that will end in miscarriage or induced abortions. Only 44.2% of miscarriages and induced abortions had a prior antenatal code. This could have resulted in underestimation of the number of pregnant women and attenuation of associations. Since antenatal visits are without information on gestational length information, we defined pregnancy start date and durations for ongoing pregnancies and early abortions based on known distributions. We chose a strict approach in the main analyses to minimize misclassification of “non pregnant” days as “pregnant”, which likely resulted in some true “pregnant” days counted as “non-pregnant” days. However, several sensitivity analyses with other assumptions of gestational lengths for these pregnancies yielded very similar results, indicating little impact on associations. Another limitation was that we could not adjust for some potential confounding factors, such as crowded living conditions, body-mass index or smoking. We were not able to look at other measures of severity such as admission to intensive-care unit due to small numbers.

In line with previous studies,^2,3,5^ our results support that pregnant women are at increased risk of severe COVID-19. However, prior studies did not compare pregnant and non-pregnant women in the general population. Among hospitalized women, others have found that pregnant women have an increased risk of intensive care and death when compared to non-pregnant women.^2,5,^ A recent meta-analysis of 123,176 non-pregnant and 10,000 pregnant women reported a higher case-fatality rate in pregnant women.^6^ As pregnant women may be more likely to be admitted to hospitals than non-pregnant women with similar symptoms, restricting studies to women hospitalized with COVID-19 may complicate interpretation of results. We found a higher risk of hospitalization when pregnant, but a similar duration of the hospital stay and slightly lower proportion with co-registrations of lower respiratory illness, compared to non-pregnant women. This may suggest that, in Norway, when hospitalized, there is no substantial difference in severity of disease in pregnant women, although more detailed data is needed to address this.

Even though several studies conclude that pregnant women are at higher risk of severe COVID-19^1^ and of adverse pregnancy outcomes in women with COVID-19,^5,17^ vaccination of pregnant women against COVID-19 is currently debated.^18-21^ COVID-19 vaccines have not been tested in pregnant women, and pregnant women are in general not recommended vaccination but to be evaluated on an individual basis.^22,23^ We found that pregnant women were not at higher risk of SARS-CoV2 infection per se, however, our results support the current evidence that there may be an increased risk of hospitalization when infected during pregnancy. Protecting pregnant women against COVID-19 is therefore important, and there is an urgent need to address vaccine safety in pregnancy.

In conclusion, in this large nationwide registry study, pregnant women were not at higher risk of SARS-CoV2 infection, but pregnancy increased the risk of specialist care and hospitalization for COVID-19 compared to non-pregnant women of the same age. Pregnant women born outside of Scandinavia were of a particular increased risk, and increased surveillance in this group is warranted. The increased risk of severe COVID-19 support the urgent need of evaluating vaccinations for pregnant women.

This research was supported by the Research Council of Norway through its Centres of Excellence funding scheme (project number 262700) and NordForsk (project number 105545). MCM works at the Medical Research Council (MRC) Integrative Epidemiology Unit at the University of Bristol which receives infrastructure funding from the UK MRC (MC_UU_00011/6). The funders had no role in the completion of the research project, the writing of the manuscript for publication, or the decision to publish the results.

## Data availability statement

Data are available by applying the Norwegian registry owners: https://helsedata.no/soknadsveiledning/.

## Data Availability

Data availability statement: Data are available by applying the Norwegian registry owners: https://helsedata.no/soknadsveiledning/.

