## Supplementary Appendix for "Pregnancy and risk of COVID-19"

This appendix has been provided by the authors to give readers additional information about their work.

**Table of contents**

**Data sources and linkages**……………………………………………………………….... 3

**Figure S1** Gestational age distribution of the first registration of antenatal codes….…………5

**Figure S2** Probability of testing in pregnant versus non-pregnant women per month………... 6

**Table S1** Codes used to define miscarriages and induced abortions………………………….. 7

**Table S2** Codes used to identify ongoing pregnancies……………………………………... 8

**Table S3** Codes used to define underlying conditions and risk groups………………………. 9

**Table S4** Risk of a positive COVID-19 test during pregnancy, excluding those who were positive 3 days before or after the end of pregnancy……….……………….…….10

**Table S5** Risk of a positive COVID-19 test in women born outside of Scandinavia……….. 11

**Table S6** Risk of specialist COVID-19 diagnoses in women born outside of Scandinavia…. 12

**Table S7** Risk of COVID-19 hospitalization in women born outside Scandinavia………..… 13

**Table S8** The probability of being tested for COVID-19 when pregnant…………….………… 14

**Table S9** The probability of being tested for COVID-19 when pregnant in women born outside Scandinavia….....………………………..………………..………15

**Details of data sources and linkages**

Data in Norwegian national health registries are registered with a personal identification number (pin), which is given all citizens in Norway for identification and public administration purposes. This pin enables linkages of individual level information across registries.

*The Emergency Preparedness Register for COVID-19*

Data in this study was provided through the Emergency preparedness register for COVID-19 (Beredt C19) administered by the Norwegian Institute of Public Health, according to the Health Preparedness Act §2-4. This registry was established in 2020 to give authorities up to date data for generating knowledge of the prevalence, causal relationships, and consequences of the COVID-19 epidemic in Norway. Beredt C19 includes information that has already been collected in the healthcare service, national health registries and administrative registers with information about the Norwegian population. The data subjects' right is safeguarded as they can contact the data controller for all different sources included in Beredt C19 in the usual way.

Through Beredt C19 we used data from the following sources:

*The National Population Register*

This register contains information on all persons in Norway. We used this registry to define the study population by including all women registered in this registry between ages 15-45 on March 1st,2020. We included dates for emigration and death to censor women correctly.

*The Medical Birth Registry of Norway (MBRN)*

The Norwegian national birth registry includes information on all pregnancies ending in gestational 12 or later from 1967 onwards. The registry is registered with the woman's pin includes information on live births, stillbirths, late miscarriages, and late induced abortions. Midwives register information on maternal background characteristics and health during pregnancy, in addition to information on pregnancy outcomes and neonatal health of offspring.

*The Norwegian Patient Registry (NPR)*

The Norwegian patient registry includes individual level information on all contacts with specialist health-care services. Information registered includes the pin, admission and discharge dates, the type of department and level of care received, in addition to diagnostic codes during the hospital stay. These discharge codes are coded according to the International Classification of Diseases version 10. We used the discharge codes to identify codes indicating the presence of a miscarriage or induced abortions. With cross- referencing against the birth registry (checking that codes were not within a registered pregnancy in the birth registry which has pregnancy ending after gestational week 12), we used the NPR registry to identify miscarriages and induced abortions that occurred before 12 completed gestational weeks. We also identified contact with specialist health-care services for suspected (ICD-10 code U07.2) or confirmed (ICD-10 code U07.1) COVID-19, lower respiratory illness during the COVID-19 hospital stay, and preexisting underlying health conditions (see Table S3)

*Norwegian Registry of Primary Health Care (KPR)*

This registry was established in 2017. Information registered includes the pin, the date, the reimbursement code for payment of services, and medical diagnoses. The diagnoses are coded according to the International Classification of Primary Care version 2. We used a selection of these codes (see Table S1) to identify miscarriages occurring before 12 gestational weeks when pregnancies were not registered in the birth or patient registry. The subgroup of miscarriages identified through the primary care registry reflect those only seen by a general practitioner that were not referred for follow up in the specialist health-care services.

*Norwegian Surveillance System for Communicable Diseases (MSIS)*
There is mandatory reporting of selected infectious diseases to this National Health register. Reporting of all COVID-19 test is mandatory, and this register contains date of testing, test results, and the personal identification number for each citizen.

*Statistics Norway (SSB)*
Administrative data is mandatorily reported to Statistics Norway. We used information from this database on household income in 2018, type of education and years of education completed by 2019.

**Figure S1. Distribution of the gestational age at first registration of codes we used to define ongoing pregnancies. These are shown for pregnancies registered in the birth registry which contains dates for pregnancy onset and gestational age.**

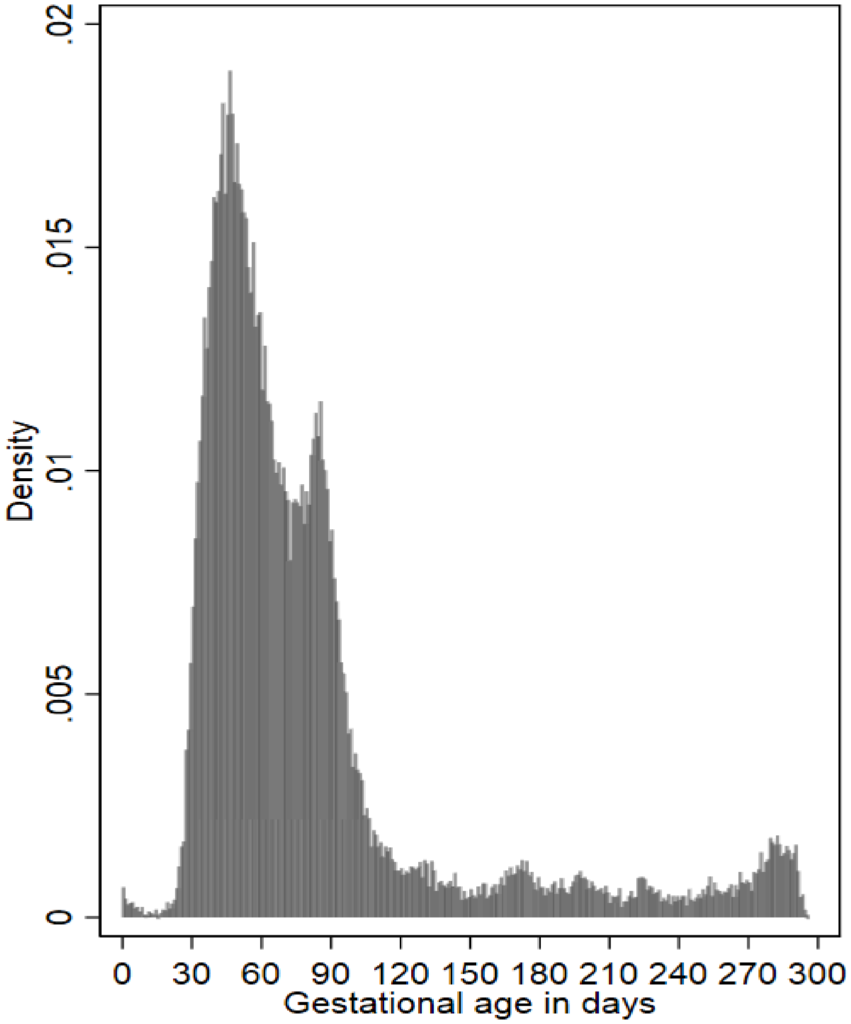

**Figure S2. The monthly Hazard ratio for being tested for SARS-CoV-2 when pregnant as compared to non-pregnant women from March 2020 to February 2021.**

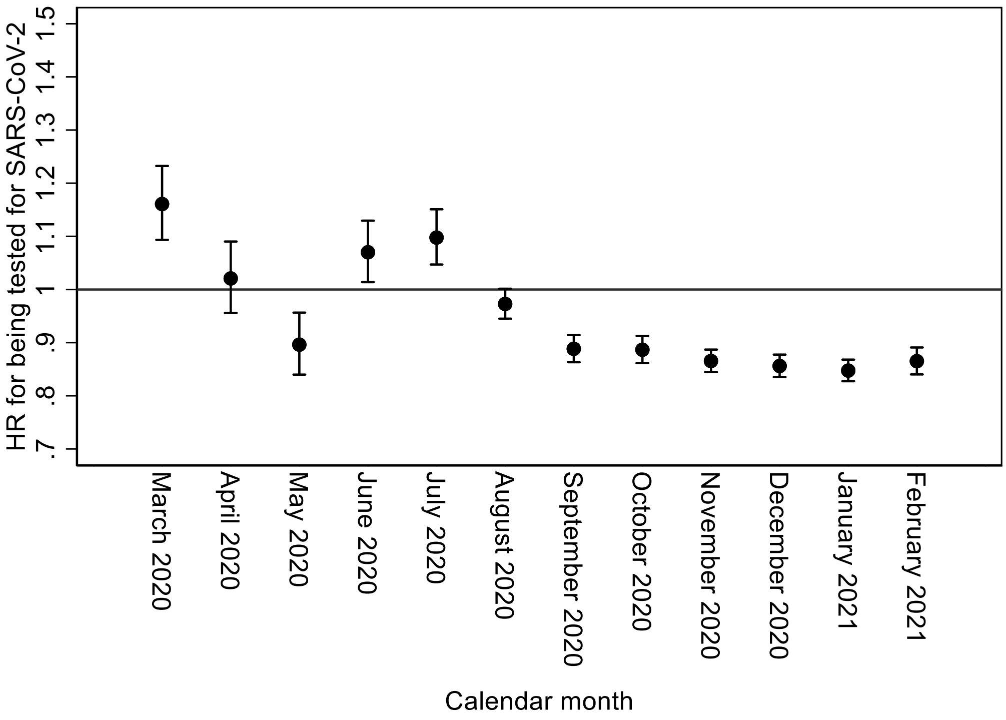

**Table S1. Diagnostic codes used to define miscarriages and induced abortions in the patient registry and general practitioner database. These were used to identify pregnancies that were not registered in the birth registry.**

| | |
|---|---|
| **ICD-10 codes used to define miscarriage in the patient registry** | |
| **O01** | Hydatidiform mole |
| **O02.0** | Blighted ovum and non-hydatidiform mole |
| **O02.1** | Missed abortion |
| **O02.8** | Other specified abnormal products of conception |
| **O02.9** | Abnormal product of conception, unspecified |
| **O03** | Spontaneous abortion |
| **O02.0** | Threatened abortion |
| **ICD-10 codes used to define induced abortions in the patient registry** | |
| **O04** | Medical abortion |
| **O05** | Other abortion |
| **O06** | Unspecified abortion |
| **ICPC-2 codes use to define miscarriage from general practitioner database** | |
| **W82** | Spontaneous abortion |

**Table S2. Diagnostic codes in the patient registry and general practitioner database used to identify ongoing pregnancies that had not yet ended and therefore were not registered in the birth registry.**

| |
|---|
| **ICD-10 codes in the patient registry** |
| O10 Pre-existing hypertension complicating pregnancy, childbirth and the puerperium |
| O11 Pre-eclampsia superimposed on chronic hypertension |
| O12 Gestational [pregnancy-induced] oedema and proteinuria without hypertension |
| O13 Gestational [pregnancy-induced] hypertension |
| O14 Pre-eclampsia |
| O15 Eclampsia |
| O16 Unspecified maternal hypertension |
| O20 Hemorrhage in early pregnancy |
| O21 Excessive vomiting in pregnancy |
| O22 Venous complications and hemorrhoids in pregnancy |
| O23 Infections of genitourinary tract in pregnancy |
| O24 Diabetes mellitus in pregnancy |
| O25 Malnutrition in pregnancy |
| O40 Polyhydramnion |
| O41 Other disorders of amniotic fluid and membranes |
| O43 Placental disorders |
| O44 Placenta praevia |
| O46 Antepartum hemorrhage,not elsewhere classified |
| O47 False labor |
| O48 Prolonged pregnancy |
| O26 Maternal care for other conditions predominantly related to pregnancy |
| O28 Abnormal findings on antenatal screening of mother |
| O29 Complications of anesthesia during pregnancy |
| O30 Multiple gestation |
| O31 Complications specific to multiple gestation |
| O32 Maternal care for known or suspected malpresentation of fetus |
| O33 Maternal care for known or suspected disproportion |
| O34 Maternal care for known or suspected abnormality of pelvic organs |
| O35 Maternal care for known or suspected fetal abnormality and damage |
| O36 Maternal care for other known or suspected fetal problems |
| **ICPC-2 codes from general practitioner database** |
| W05 Pregnancy vomiting/nausea |
| W81 Toxemia of pregnancy |
| W84 Pregnancy high risk |
| W85 Gestational diabetes |
| W03 Antepartum bleeding |
| W21 Concern about body image related to pregnancy |
| W28 Limited function/disability (W) |
| W71 Other infection complicating pregnancy/puerperium |
| W72 Malignant neoplasm related to pregnancy |
| W73 Benign/unspecified neoplasm related to pregnancy |
| W75 Injury complicating pregnancy |
| W76 Congenital anomaly complicating pregnancy |
| W79 Unwanted pregnancy |
| W781 Prenatal follow-up |
| W78 Pregnancy |

**Table S3. Codes used to define underlying conditions and risk groups. Codes from January 2017 through February 2021 were included.**

| Risk condition | ICD-10 | ICPC2 |
|---|---|---|
| Organ transplant | Z94.0, Z94.1, Z94.2, Z94.3, Z94.4, Z94.8 | |
| Neurological disorders | G1, G20, G21, G23, G24, G40.5, G61.0, G70, G71, G80.0, G80.2, G80.3, F72, F73, F84.0, F84.1, Q05.0, Q05.1, Q05.2, Q05.3, Q05.04, Q05.5, Q05.6, Q90 | |
| Kidney failure | N18.3, N18.4, N18.5 | |
| Diabetes | E10, E11, E12, E13, E14 | T89, T90 |
| Chronic lung disease | J41, J42, J43, J44, J45, J46, J47, J84, J98, E84 | R95, R96 |
| Hematological cancer | C81, C82, C83, C84, C85, C86, C87, C88, C89, C90, C91, C92, C93, C94, C95, C96, D45, D45, D47 | |
| Other Cancers | C0, C1, C2, C3, C4, C5, C6, C7, C80, D32, D33, D35.2, D35.3, D35.4, D42, D43, D44.2, D44.3, D44.4, | |
| Immunodeficiency | D80, D81, D82, D83, D84 | |
| Reduced immune function | G35, M05, M08, M06, M07, M09, M13, M14, K50, K51 | |
| Cardiovascular disorders | I05, I06, I07, I08, I09, I2, I31, I32, I34, I35, I36, I37, I39, I40, I41, I42, I43, I46, I48, I49, I50 | K74, K75, K76, K77, K78, K82, K83, K87 |
| Cerebrovascular disease | I60, I61, I62, I63, I64, I69.1, I69.2, I69.3, I69.4, I69.8, I69.0 | K90, K91 |

**Table S4. Hazard ratios of a positive SARS-CoV-2 test during pregnancy when excluding women who were positive within 3 days before or after the end of pregnancy, among 1,033,647 women between 15 and 45 years of age.**

| Pregnancy status | Follow-up time in days | No. of positive tests | Hazard Ratio (95% CI) | |
|---|---|---|---|---|
| | | | Without adjustment | With Adjustment* |
| Not pregnant | 356,378,916 | 16,364 | 1.00 | 1.00 |
| Pregnant | 15,475,225 | 677 | 0.93 (0.86-1.01) | 0.95 (0.88-1.03) |
| 1st trimester | 5,451,393 | 253 | 0.96 (0.85-1.09) | 0.97 (0.86-1.11) |
| 2nd trimester | 5,785,453 | 260 | 0.95 (0.84-1.08) | 0.97 (0.86-1.10) |
| 3rd trimester | 4,238,379 | 164 | 0.87 (0.74-1.01) | 0.88 (0.76-1.03) |

*Adjusted for age as a linear and squared term, country of birth, marital status, education, household income, diabetes, cerebrovascular disease, other cardiovascular disorders, immune-deficiency, chronic lung disease, reduced immune function, neurological disorders, kidney failure, organ transplant, hematological cancer, and other types of cancer.

**Table S5. Hazard ratios of a positive SARS-CoV-2 test according to being born outside of Scandinavia and being pregnant among 1,030,421* women between 15 and 45 years of age in Norway.**

| **Born in Scandinavia** | **Pregnancy status** | **Follow-up time in days** | **No. Positive tests** | **Hazard Ratio (95% CI)** Without Adjustment | With Adjustment† |
|---|---|---|---|---|---|
| Yes | Not pregnant | 275,928,056 | 10,500 | 1.00 | 1.00 |
| Yes | Pregnant | 11,495,462 | 358 | 0.80 (0.72-0.89) | 0.85 (0.76-0.94) |
| No | Not pregnant | 79,321,894 | 5837 | 1.95 (1.88-2.01) | 2.13 (2.05-2.21) |
| No | Pregnant | 3,935,918 | 349 | 2.34 (2.11-2.60) | 2.57 (2.31-2.87) |
| No | Not pregnant | 79,321,894 | 5837 | 1.00 | 1.00 |
| No | Pregnant | 3,935,918 | 349 | 1.20 (1.08-1.34) | 1.24 (1.11-1.38) |
| Yes | Pregnant | 11,495,462 | 358 | 1.00 | 1.00 |
| No | Pregnant | 3,935,918 | 349 | 2.94 (2.53-3.40) | 2.37 (1.98-2.84) |

*Excluding one person who tested positive before March 1st, 2020, and 3277 women with unknown country of birth.
†Adjusted for age as a linear and squared term, country of birth, marital status, education, household income, diabetes, cerebrovascular disease, other cardiovascular disorders, immune-deficiency, chronic lung disease, reduced immune function, neurological disorders, kidney failure, organ transplant, hematological cancer, and other types of cancer.

**Table S6. Risk of a specialist care diagnosis of COVID-19 (event) according to being born outside of Scandinavia and being pregnant among 1,030,419* women between 15 and 45 years of age.**

| Born in Scandinavia | Pregnancy status | Follow-up time in days | All events | | | Excluding concurrent events† | |
|---|---|---|---|---|---|---|---|
| | | | No. of events | Hazard Ratio (95% CI) | | No. of events | Hazard Ratio (95% CI) |
| | | | | Without Adjustment | With Adjustment‡ | | With Adjustment‡ |
| Yes | Not pregnant | 277,034,136 | 638 | 1.00 | 1.00 | 638 | 1.00 |
| Yes | Pregnant | 11,534,155 | 76 | 2.84 (2.24-3.60) | 2.66 (2.09-3.39) | 51 | 1.80 (1.35-2.41) |
| No | Not pregnant | 79,893,467 | 260 | 1.41 (1.22-1.63) | 1.44 (1.23-1.69) | 260 | 1.45 (1.24-1.70) |
| No | Pregnant | 3,964,970 | 68 | 7.44 (5.80-9.56) | 7.50 (5.76-9.77) | 36 | 4.05 (2.86-5.73) |
| No | Not pregnant | 79,893,467 | 260 | 1.00 | 1.00 | 260 | 1.00 |
| No | Pregnant | 3,964,970 | 68 | 5.27 (4.03-6.88) | 6.07 (4.61-7.98) | 36 | 3.28 (2.29-4.68) |
| Yes | Pregnant | 11,534,155 | 76 | 1.00 | 1.00 | 51 | 1.00 |
| No | Pregnant | 3,964,970 | 68 | 2.61 (1.88-3.62) | 2.37 (1.63-3.46) | 36 | 1.80 (1.11-2.94) |

*Excluded three people in contact with specialist health-care diagnose of COVID-19 before March 1st, 2020 and 3277 women with unknown country of birth.

†Excluding concurrent events where the end of pregnancy occurred within the hospital stay for COVID-19.

‡Adjusted for age as a linear and squared term, country of birth, marital status, education, household income, diabetes, cerebrovascular disease, other cardiovascular disorders, immune-deficiency, chronic lung disease, reduced immune function, neurological disorders, kidney failure, organ transplant, hematological cancer, and other types of cancer.

**Table S7. Risk of hospitalization (event) for confirmed COVID-19 according to being born outside of Scandinavia and being pregnant among 1,030,422 women[*] between 15 and 45 years of age.**

| Born in Scandinavia | Pregnancy status | Follow-up time in days | All events | | | Excluding concurrent events[†] | |
|---|---|---|---|---|---|---|---|
| | | | No. of events | Hazard Ratio (95% CI) | | No. of events | Hazard Ratio (95% CI) |
| | | | | Without Adjustment | With Adjustment[‡] | | With Adjustment[‡] |
| Yes | Not pregnant | 277,121,220 | 155 | 1.00 | 1.00 | 155 | 1.00 |
| Yes | Pregnant | 11,541,027 | 17 | 2.61 (1.58-4.30) | 3.11 (1.89-5.14) | 6 | 1.15 (0.51-2.60) |
| No | Not pregnant | 79,916,083 | 132 | 2.96 (2.34-3.73) | 2.54 (1.98-3.26) | 132 | 2.59 (2.01-3.33) |
| No | Pregnant | 3,968,676 | 36 | 16.24 (11.30-23.34) | 16.02 (10.90-23.55) | 18 | 8.47 (5.10-14.07) |
| No | Not pregnant | 79,916,083 | 132 | 1.00 | 1.00 | 132 | 1.00 |
| No | Pregnant | 3,968,676 | 36 | 5.49 (3.79-7.94) | 6.94 (4.79-10.06) | 18 | 3.63 (2.19-5.60) |
| Yes | Pregnant | 11,541,027 | 17 | 1.00 | 1.00 | 6 | 1.00 |
| No | Pregnant | 3,968,676 | 36 | 6.25 (3.51-11.12) | 4.72 (2.51-8.87) | 18 | 6.12 (2.26-16.57) |

[*] Excluding 3277 women with unknown country of birth.

[†] Excluding concurrent events where the end of pregnancy occurred within the hospital stay for COVID-19.

[‡] Adjusted for age as a linear and squared term, country of birth, marital status, education, household income, diabetes, cerebrovascular disease, other cardiovascular disorders, immune-deficiency, chronic lung disease, reduced immune function, neurological disorders, kidney failure, organ transplant, hematological cancer, and other types of cancer.

**Table S8. The likelihood of being tested for SARS-CoV-2 while pregnant among 1,033,699 women between 15 and 45 years of age.**

| Follow-up period | Pregnancy status | No. of days of follow-up | All tests | | | Excluding tests 3 days before or after end of pregnancy | |
|---|---|---|---|---|---|---|---|
| | | | No. of tests | Hazard Ratio (95% CI) | | No. of tests | Hazard Ratio (95% CI) |
| | | | | Without Adjustment | With Adjustment* | | With Adjustment* |
| **Complete follow-up**† | Not pregnant | 358,589,148 | 1,116,251 | 1.00 | 1.00 | 1,116,115 | 1.00 |
| | Pregnant | 15,567,021 | 45,488 | 0.93 (0.92-0.94) | 0.90 (0.88-0.91) | 42,593 | 0.84 (0.83-0.85) |
| | 1st trimester | 5,484,461 | 18,059 | 1.03 (1.01-1.04) | 0.99 (0.97-1.00) | 17,124 | 0.94 (0.92-0.95) |
| | 2nd trimester | 5,820,579 | 16,544 | 0.90 (0.89-0.92) | 0.86 (0.85-0.88) | 16,243 | 0.85 (0.83-0.86) |
| | 3rd trimester | 4,261,981 | 10,885 | 0.85 (0.83-0.87) | 0.81 (0.80-0.83) | 9226 | 0.69 (0.67-0.71) |
| **Wave 1**‡ | Not pregnant | 119,622,006 | 92,707 | 1.00 | 1.00 | 92,681 | 1.00 |
| | Pregnant | 5,205,781 | 4287 | 1.06 (1.03-1.10) | 0.95 (0.92-0.98) | 3,949 | 0.87 (0.85-0.90) |
| | 1st trimester | 1,749,424 | 1620 | 1.19 (1.13-1.26) | 1.08 (1.02-1.14) | 1531 | 1.02 (0.97-1.08) |
| | 2nd trimester | 1,943,922 | 1411 | 0.94 (0.89-1.00) | 0.84 (0.79-0.88) | 1404 | 0.84 (0.79-0.88) |
| | 3rd trimester | 1,512,435 | 1256 | 1.07 (1.01-1.13) | 0.93 (0.88-0.99) | 1014 | 0.76 (0.71-0.81) |
| **Wave 2**§ | Not pregnant | 238,967,142 | 1,023,544 | 1.00 | 1.00 | 1,023,434 | 1.00 |
| | Pregnant | 10,361,240 | 41,201 | 0.92 (0.91-0.93) | 0.90 (0.88-0.90) | 38,644 | 0.83 (0.82-0.84) |
| | 1st trimester | 3,735,037 | 16,439 | 1.01 (1.00-1.03) | 0.98 (0.96-1.00) | 15,593 | 0.93 (0.91-0.95) |
| | 2nd trimester | 3,876,657 | 15,133 | 0.90 (0.88-0.92) | 0.86 (0.85-0.88) | 14,839 | 0.85 (0.83-0.86) |
| | 3rd trimester | 2,749,546 | 9629 | 0.82 (0.81-0.84) | 0.80 (0.78-0.82) | 8212 | 0.68 (0.67-0.70) |

* Adjusted for age as a linear and squared term, country of birth, marital status, education, household income, diabetes, cerebrovascular disease, other cardiovascular disorders, immune-deficiency, chronic lung disease, reduced immune function, neurological disorders, kidney failure, organ transplant, hematological cancer, and other types of cancer.

† March 1st, 2020 to February 28th, 2021

‡ March 1st, 2020 to June 30th, 2020

§ July 1st, 2020 to February 28th, 2021

**Table S9. The likelihood of being tested for SARS-CoV-2 according to being born outside of Scandinavia and being pregnant among 1,030,422 women* between 15 and 45 years of age in Norway.**

| Born in Scandinavia | Pregnancy status | Follow-up time in days | No. of tests | Hazard Ratio (95% CI) | |
|---|---|---|---|---|---|
| | | | | Without Adjustment | With Adjustment† |
| Yes | Not pregnant | 277,440,147 | 933,843 | 1.00 | 1.00 |
| Yes | Pregnant | 11,545,059 | 36,976 | 0.94 (0.93-0.95) | 0.89 (0.88-0.90) |
| No | Not pregnant | 80,012,624 | 179,091 | 0.67 (0.66-0.67) | 0.78 (0.77-0.78) |
| No | Pregnant | 3,971,779 | 8355 | 0.63 (0.61-0.65) | 0.72 (0.70-0.74) |
| No | Not pregnant | 80,012,624 | 179,091 | 1.00 | 1.00 |
| No | Pregnant | 3,971,779 | 8355 | 0.95 (0.92-0.97) | 0.98 (0.96-1.01) |
| No | Not pregnant | 11,545,059 | 36,976 | 1.00 | 1.00 |
| No | Pregnant | 3,971,779 | 8355 | 0.67 (0.65-0.69) | 0.76 (0.74-0.79) |

* Excluding 3277 women with unknown country of birth.
† Adjusted for age as a linear and squared term, country of birth, marital status, education, household income, diabetes, cerebrovascular disease, other cardiovascular disorders, immune-deficiency, chronic lung disease, reduced immune function, neurological disorders, kidney failure, organ transplant, hematological cancer, and other types of cancer.
